# Prevalence of Modifiable Risk Factors for Cardiovascular Disease among School-going Children and Adolescents in Eldoret, Kenya

**DOI:** 10.1101/2023.08.24.23294556

**Authors:** Karani Magutah, Grace Wambura Mbuthia, Gilbert Osengo, Dancun Odhiambo, Rebecca Meiring

## Abstract

Cardiovascular disease (CVD) prevalence in Kenya is rising. Overweight, pre-hypertension and physical inactivity in younger ages is contributory. These risk factors are inadequately documented among Kenyan children and adolescents, hampering CVD prevention.

This cross-sectional study randomly sampled 384 school children and adolescents. After obtaining assent and consent, global physical activity (PA) questionnaire was used to assess PA. Body mass index (BMI), Waist-Hip Ratio (WHR) and Waist-Height Ratio (WHtR) were determined. Blood pressure (BP) was also measured.

Participants were 14.6±2.7 years, and 62.6% were female. Overall BMI was 19.8±3.9 kg/m^2^ with 8% having ≥25.0 kg/m^2^, 87% of whom were in secondary schools. Using SBP, 27.9% were at risk of CVD. For age ≥13 years old, males and females proportions were 42.5% and 20% respectively while that for <13 years old, it was 26.5% and 27% respectively. For DBP, 12.8% had elevated-to-hypertensive BP. For ages ≥ 13 years old, this was 13.2% and 8.3% for males and females respectively and for the <13 years old, the respective percentages were 11.8 and 25.4. Combining SBP and DBP, 8.1% of participants, mostly males, had elevated-to-hypertensive BP. Thirty-one percent of boys and 15.6% of girls were at CVD risk using respective WHR cutoffs of 0.90 and 0.85 (males, 0.93±0.02; females, 0.89±0.03). For WHtR, 39.6% of boys had values >0.463 cut-off (0.493±0.02), with 52.6% in secondary schools against 32.4% for girls having >0.469 cut-off (0.517±0.05), 69.7% being in secondary schools. Overall, 45% of participants were sports-inactive and 77.2% did minimal PA.

Among school-going children and adolescents in Eldoret, Kenya, prevalence of CVD risk-factors was high especially among boys and in high schools. Large proportions had elevated BP, BMI, WHR and WHtR, and, further, were sedentary, posing high CVD risk. Lifestyle interventions to mitigate this public health concern among children and adolescent are urgently needed.

## Introduction

The prevalence of cardiovascular disease (CVD) is increasing in sub-Saharan Africa, a region previously thought to suffer mainly from infectious diseases only. The recently observed trend of increasing sedentary lifestyles in Kenya (1, 2) is likely to contribute to the increased prevalence of CVD and associated mortalities in Africa, and will likely result in a significant economic burden to individuals and the country at large (3). Without intervention, it takes only four years for 20-33% of pre-hypertensive individuals to develop CVD (4). This statistic is a concern in Kenya, as it would further burden the economy that already attracts a high per capita cost in the management of CVD (5). The majority of the Kenyan population is poor, living on <1 USD a day (6).

Much research has identified risk factors for CVD amongst adults (7–9) globally. The prevalence of some risk factors, such as pre-hypertension, is as high as 47% in Kenyan adults (51% for males; 46% for females) (10). While CVD more commonly occurs in middle-age to older adults, certain modifiable risk factors during childhood and adolescence lead to an increased risk of developing CVD as an adult (11–13). These risk factors for CVD include overweight and obesity, pre- hypertension, physical inactivity and high sedentary behaviour (11, 12, 14–16). Prevalence studies among children and adolescents have however been minimal and the prevalence of modifiable risk factors in these cohorts is not confirmed in Kenya. Recently, one study of primary school children showed 9% had elevated BP while 33% had stage 1 hypertension. In addition, higher BP was associated with higher body mass. (17). While elevated BP and hypertension in children and adolescents continues raising concern not only in both the developed and developing economies (18, 19) with a recent systematic review showing prevalence of prehypertension among cohorts in Africa ranging from 2.5-34% (20), we have minimal data from Kenya. In the sub-Saharan region, the prevalence of prehypertension and hypertension in South African primary school children was recently documented ranging 5.2-18.5% (21). Elevated BP, Prehypertension and hypertension stands at 18.1%, 9.6% and 8.5% respectively in Tanzania (22). Uncontrolled, these risk factors will contribute significantly to the development of CVD in adulthood (23).

Interventions such as physical activity (PA) reduce cardiovascular-related morbidity risk and/or modification of the disease for those already affected (24). When done early in life, such involvement in PA may become a lifestyle carried into adulthood, helping prevent CVD occurrence in those more at risk (15, 16, 25, 26). School programs and screen-time at home have however contributed to behaviours that are associated with inactivity or sedentariness posing a risk for and the development of enhanced CVD risk factors among school-going children and adolescents (27, 28). We are starting to observe similar trends in Kenya coming at a time when sedentary behavior reduction is now included as an independent factor for consideration in the quest for optimal health (29).

Early screening can help identify at-risk children and adolescents for timely interventions to reduce the occurrence of CVD risk factors. There are realistic and affordable screening approaches available such as the measurements of body composition (BC) and BP as well as methods for the assessment of physical activity and sedentary behaviour through which we are able to find those at a greater risk as well as those already hypertensive. From such measurements we could have an objective documentation of the prevalence of risk factors associated with CVD among children and adolescents in Kenya.

A clear picture of the current prevalence of risk factors associated with the now or future CVD development among Kenyan children and adolescents could help give clarity and, further, evidence needed for designing early interventions necessary to mitigate the problem.

## Methods

### Design and Setting

This was a cross-sectional observational study conducted in Eldoret, Kenya.

### Study population

Participants were school going children and adolescents in the upper classes of primary school and high schools in Kenya. This provided a more comparable age bracket since they have similar cut-offs for various variables studied.

### Sampling Procedure

We studied 384 participants from primary and secondary schools in the metropolitan town of Eldoret, Kenya. The sample size was determined using a CVD risk factors’ assumption of 0.5 proportion that would provide the largest possible sample given unavailability of Kenyan data for physically inactive / sedentary children and adolescents, or data for other risk factors associated with future development of CVD from Kenya. Participants were drawn from public and private primary and secondary schools. To ease comparisons at analysis, the last four classes of primary school and the four classes of secondary school (as is the system in Kenya) were selected to be invited into the study. The selection for the primary and secondary public and private schools for study was random, picking random papers each with a name of a school from a list of all schools in the metropolis. The numbers studied per school were proportionately allocated based on the student population at each school. To ensure equal chances for participation, selection of the individual participants utilized computer generated random numbers from the class registers.

### Eligibility

Upper primary and secondary school-going children and adolescents within Eldoret were included. Participants excluded were those with physical impairments which could compromise participation in PA as this has been associated with higher CVD risk. These physical challenges are not modifiable risk factors. We also excluded participants with cognitive inabilities that would affect administration of the study tool.

### Ethical Consideration

This study was approved by the Moi Teaching and Referral Hospital / Moi University Institutional Research Ethics Committee (MTRH/MU IREC) on 22^nd^ November 2022 (approval number FAN: 0004291) and obtained research permit from the National Commission for Science, Technology and Innovation (NACOSTI) on 1^st^ March 2023 (permit Ref number 839919). After having the study protocol fully verbally explained, children and adolescents provided written assent and their parents / guardians, further, provided written informed consent.

### Assessments

The first participant was recruited on 6^th^ March 2023 and engagement with the last participant was on 28^th^ April 2023. The global PA questionnaire (GPAQ) was used to assess the level of physical activity and sedentary behaviour. Weight and height were measured using calibrated weighing machine (CAMRY Mechanical scale, BR9012, Shanghai, China) and a tape measure with participants wearing light clothing and no shoes. Waist circumference (WC) was measured with a tape measure, as the diameter horizontally all round just above the iliac crest and hip circumference (HC) as the widest diameter across the buttocks. All measurements were made in centimeters. BP was measured using Omron M2 Basic (HEM-7120-E) automatic BP monitor (Omron Healthcare Co. Ltd, Kyoto, Japan). Two BP measurements were taken 2 minutes apart following a 5 minutes’ rest in a sitting position and the average was recorded

### Data Management and Analysis

Data were analyzed using STATA v.13 at univariate (proportions, means and standard deviation for BP, WC, Waist-Hip Ratio (WHR), Waist-Height Ratio (WHtR) and body mass index (BMI). Analyses for PA engagement was based on proportions that were either active or sedentary. The WHR, WHtR and BMI were determined from the measurements of WC, HC, weight and height. All these were thereafter re-analyzed based on applicable cut-offs to identify participants’ risk levels. Participants with values above their set cut- offs for all or either of WHtR (0.463 for boys and 0.469 for girls) (30–32), WHR (≥0.90 (male) and ≥0.85 (female) (33), BMI (A 95 percentile of adult values applied amongst <13 year old and a similar percentile used for BMI categorization for all participants’ age range) (34, 35) and BP (≥120/80 for ages ≥13 years) (35) were considered at risk for CVD.

## Results

There were 145 (37.4%) female and 243 (62.6%) male participants. The average age of participants was 14.6±2.7 years. Those ≥13 years (n=285) had mean age 15.6± 2.2 years while those <13 years (n=99) had 11.4±0.8 mean age. Thirty four percent (33.8%), 13.1%, 30.5% and 22.6% were drawn from public secondary, private secondary, private primary and public primary schools respectively. The class distributions’ for participants ranged from 8.5% for the lowest representation (grade 5) to 20.5% for the highest (class 8) using the Kenyan education system.

Eight percent (7.6%, n=29) of the participants had hypertensive pressure ≥130 mmHg (≥124 mmHg for <13 year old) using SBP, 18.5% had prehypertensive ranges 120-129 mmHg (114- 123mmHg for < 13 year old) and 73.9% had normal SBP for their ages. For males and females respectively, 25% (n=35) and 14.8% (n=36) were pre-hypertensive while 9.3% (n=13) and 6.6% (n=16) had hypertensive SBPs. Using DBP, 7.3% (n=28) of participants were in elevated BP range of 80-89 mmHg (≥76-85 mmHg) for <13 years old) and 1.3% (n=5) were already hypertensive. For males and females respectively, 7.9% (n=11) and 7% (n=17) had prehypertensive pressures, with 0.7% (n=1) and 1% (n=4) having hypertensive DBPs ≥90 mmHg (≥86 mmHg for < 13 year old). Combining both SBP and DBP, 8.1% of participants (n=31) accounted for by a 9.3% (n=13) among males and 7.4% among females (n=18) had elevated to hypertensive BPs. A chi-squared test showed that overall, males had higher SBP compared to females (χ^2^ = 9.81; p = 0.007) and that those from private schools had higher SBP when compared to those from the private (χ^2^ = 15.54; p = <0.001). The various means for the cardiovascular and body composition variables for the 144 males and 243 females are shown in table 1.

**Table 1:**
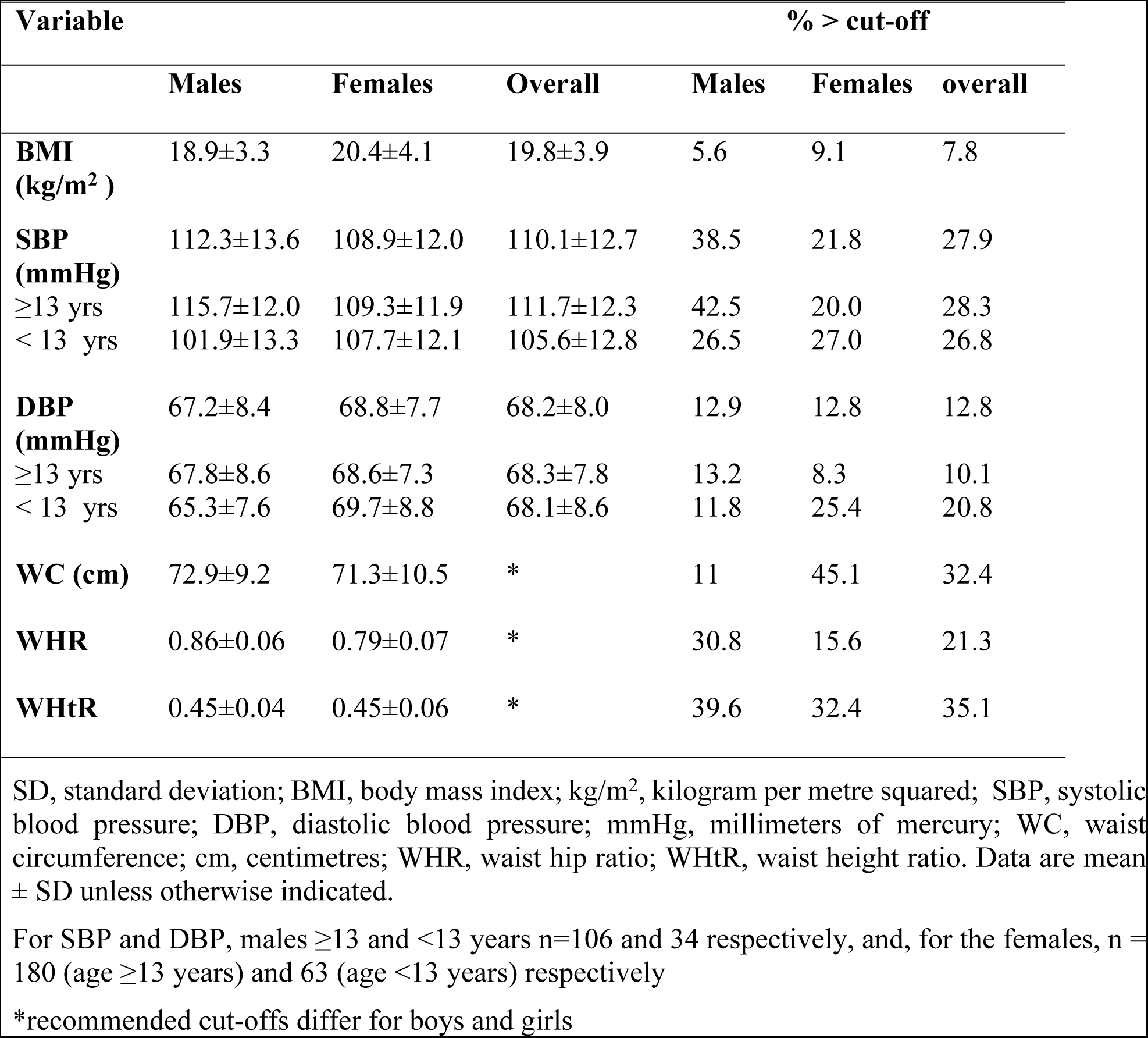
Cardiovascular and body composition means and proportions.

Six percent (6.2%) of participants (N=387) had a BMI of 25–<30 and 1.6% were at ≥30. Disaggregating to males (N=144) and females (N=243), 5.6% versus 6.6% were in the 25–<30 BMI range respectively. While 2.5% females had BMI ≥30, no males had such. Being female was associated with higher BMI (χ^2^ = 11.26; p = 0.01). Eight seven percent (86.6%) of those with BMI ≥25.0 were in secondary schools and which was associated with higher BMI (χ^2^ = 102.54; p < 0.001), and 53% came from public schools.

The mean (SD) WC for males was 72.9±9.2 centimeters and 71.3±10.5 centimeters for females. Using proposed ≥84.6 cm for boys and ≥72 cm for girls (given as 90^th^ percentile of African adult WC value ≥ 94 and 80 cm in males and females respectively) as cut-offs for WC corresponding with high CVD risk (36), 32.4% of participants (11% of boys, WC 89.8± 4.9; 45.1% of girls, WC 79.6±9.1) had higher-than-recommended values. The WHR was 0.86±0.06 and 0.79±0.07 for boys and girls respectively. Using respective WHR cut-offs of 0.90 and 0.85 for males and females, 30.8% of boys were at CVD risk (WHR 0.93±0.02) while the girls at risk were 15.6% (mean 0.89±0.03). For the boys (n=45), 42.2% were in secondary schools but for the girls (n=38), half each were from primary and secondary schools. For WHtR, it was 0.45±0.04 and 0.45±0.06 for boys and girls. For the boys, 39.6% had their WHtR above 0.463 cut-off (mean 0.493±0.02). Distribution of those with WHtR above cut-off (n=57) was 29.8% from private secondary schools, 26.3% from private primary schools, 22.8% from public secondary schools, and, 21.1% from public primary schools. Overall, 52.6% came from secondary schools while the rest were in primary. For the girls, those above respective 0.469 cut-off were 32.4% (mean 0.517±0.05). Distribution of the girls at-risk using WHtR (n=79) had 16.5% from private secondary schools, 21.5% from private primary schools, 53.2% from public secondary schools and 8.9% from public primary schools. Overall, 69.6% of girls with at-risk WHtR were in high schools compared to 30.4% in primary schools.

About 45% of the participants did not participate in any form of planned exercise activities (46.8% among males, 44.2% among females). Outside planned exercises, 77.2% (78.9% male; 76.3% female) did not participate in appreciable daily moderate or vigorous intensity PA at home or school. Distribution across schools for this and related cardiovascular and body composition variables is shown in Table 2.

**Table 2:** Percentage of participants not meeting expected levels in various variables per school category.

| **Variable** | **% of total in**<br>**PuHS (n=131)** | **% of total in**<br>**PrHS (n=51)** | **% of total in**<br>**PuPS**<br>**(n=119)** | **% of total in**<br>**PrPS (n=88)** |
| --- | --- | --- | --- | --- |
| **BMI ($\geq 25$)** | 12.2 | 19.6 | 0 | 3.4 |
| **SBP** ($\geq 13$ years, $\geq 120$mmHg; $<13$ year old, $\geq 114$ mmHg) | 29.7 | 37.2 | 22.7 | 23.8 |
| **DBP** ($\geq 13$ years, $\geq 80$mmHg; $<13$ year old, $\geq 76$ mmHg) | 10.9 | 7.8 | 11.7 | 13.6 |
| **WC** |  |  |  |  |
| Males ($\geq 84.6$ cm) | 17.1 | 25.9 | 0 | 5.4 |
| Females ($\geq 72$ cm) | 63.9 | 75 | 11.7 | 38.1 |
| **WHR** |  |  |  |  |
| Males ($\geq 0.9$) | 20 | 40.7 | 46.4 | 23.2 |
| Females ($\geq 0.85$) | 12.4 | 29.2 | 13.3 | 14.3 |
| **WHtR** |  |  |  |  |
| Males ($\geq 0.463$) | 40 | 63 | 46.4 | 26.8 |
| Females ($\geq 0.469$) | 43.3 | 54.2 | 11.7 | 27.0 |
| **Exercises** (non-participating proportion) | 44.4 | 69.4 | 20.6 | 54.2 |
| **PA** (non-participating proportion) | 49.2 | 83.7 | 93.2 | 92.4 |

PuHS, Public high school; PrHS, private high school; PuPS, public primary school; PrPS, private primary school; BMI, body mass index; SBP, systolic blood pressure; DBP, diastolic blood pressure; WC, waist circumference; WHR, waist hip ratio; WHtR, waist height ratio; PA, physical activity.

## Discussion

Participants in the current study were in mid-teens. This age has been observed to have significant modifiable CVD risk factors that include overweight and obesity, pre-hypertension, physical inactivity and high sedentary behaviours. These risk factors are more common later during the middle ages and increase through older adulthood. When they start early in teen ages, the likelihood of enhanced CVD risk in adulthood is increased (11, 12, 14).

Almost 1 in 4 participants had elevated-to-hypertensive SBPs for their ages. In every 3 males, one had elevated-to-hypertensive SBPs and there were almost twice as many affected as the females. Using DBP, about 8% were in elevated-to-hypertensive ranges and the males were still more affected. Combining both SBP and DBP also showed that males are much more affected compared to the females in our current study. A similar trend where males are more affected than females has been shown before among adults aged ≥18 years in Kenya, and, more comparatively age-wise, in neighbouring Uganda and Tanzania (10, 37, 38). While previously adult data showed up to 47% of Kenyan adults are prehypertensive, the current study shows slightly lower proportions although it involved younger participants. The at-risk proportions in the current study are within ranges found regionally in the sub-Saharan Africa and the larger continent (20, 37–39). With previous observation that up to 30% of children and adolescents aged ≥12 years who have prehypertension develop CVD later in life (15, 16), the high proportions with prehypertension in the current study may need to be addressed. There were mixed results for the mean SBP and DBP values for the males and females but higher SBP mainly affected the males. While recent works locally and regionally, just like the current study show fairly high proportions with prehypertension and hypertensive BPs among primary school children (17, 20, 22, 37–39), what the current study adds is that those in private schools had higher proportions with elevated-to-hypertensive SBPs. Further, the current study adds that those in secondary schools are more affected with about 1 in 3 having prehypertensive-to-hypertensive BPs, and, more than twice as affected as their counterparts in respective primary school category. The differences in proportions were not marked for DBP, although those from high schools were higher. The findings compares to those of a study done in Congo that showed increased odds of pre-hypertensive among school children in secondary and private schools compared to those in primary and public schools respectively(40).

Although most participants had BMI within acceptable ranges, four in every ten, and mostly boys, were underweight. Importantly given the focus of the study, appreciable proportions had overweight-to-obese BMIs, and the females were most affected. Participants from secondary schools were especially affected, probably because they were older, and public schools had a slight edge over the private. Similar findings were documented in a metaanalysis of prevalence of overweight and obesity among African primary school learners where overweight and obesity estimates were higher in private compared to public schools. This could point to the difference in social economic status and thus lifestyle behaviors of the families where these children come from. For WC, while the mean WC appeared within acceptable range for both sexes based on age (36), a third of all participants had higher-than-recommended values. Girls had four times more proportions affected, with just under a half of all females having higher than acceptable WC. It is safe to say that overall, by use of WC alone, 1 in 3 participants from the current study had central obesity and therefore enhanced CVD risk. Using respective WHR cut-offs however showed males having twice as much proportions affected compared to females and about half of such boys and girls were in both school categories. The WHtR, also showed males as having larger proportions at risk, with 2 in every 5 having WHtR >0.463. The majority of these were drawn from both private and secondary schools. For females, 1 in 3 had values >0.469 cut-off but here, although those from public secondary schools contributed the bulk overall, their proportions per school category remained, like for all other BC variables, lower compared to private schools. For all BC variables, proportions for those at higher risk were much more from high than primary schools, and, mostly, among the private.

While exercise and PA are known to reduce CVD risks (24), almost half of the current study participants did not participate in any form of planned moderate and/or vigorous intensity exercises. The participation was poorer among boys compared to girls. Similarly for PA at home or in school, the males fared slightly worse for those with minimal-to-no PA involvement. Overall, almost 4 in 5 participants were sedentary using this criterion, an exact observation first made a decade ago but among older adults (1), and, also, more recently in a study following COVID-19 associated lifestyle changes that showed little progress has occurred in exercise and PA participation in Eldoret, Kenya (2). If something is not done and these children and adolescents take a sedentary lifestyle of minimal PA involvement into their adult lives, their CVD risk could end up higher. Distribution across schools showed that higher proportions of children and adolescents from private schools did not participate in exercises and PA, and that those in high schools for both categories had lesser engagement in such planned exercises. It has previously been observed that school programs and screen-time contribute the inactivity being observed among school children and adolescents (27, 28), and although the current study did not seek this association, it is likely this contributes to the minimal exercises and PA participation observed here.

### Limitations

We used self-reports for PA and exercise data and this subjective data may not only have introduced recall bias but also compromised quality. Given that this was a cross-sectional study and, further, data was collected during school sessions made this inevitable. Data for the BC were collected by 2 research assistants and although they had been trained together, there may have been individual differences in the accuracy of the measurements taken. Further, BP measurements were done once-off in a single session. These may have affected the results.

## Conclusion

Based on the BC, BP and PA screening used in the current study, school-going children and adolescents in Eldoret, Kenya had high prevalence of CVD risk-factors. This was especially higher among boys and in high schools. Large proportions had elevated BPs, BMI, WHR and WHtR, and, further, were sedentary, posing high CVD risk. Lifestyle interventions to mitigate this public health concern and that target on children and adolescent are urgently needed.

## Financial Disclosure

KM was supported by the Consortium for Advanced Research Training in Africa (CARTA). CARTA is jointly led by the African Population and Health Research Center and the University of the Witwatersrand and funded by the Carnegie Corporation of New York (Grant No. G-19- 57145), Sida (Grant No:54100113), Uppsala Monitoring Center, Norwegian Agency for Development Cooperation (Norad), and by the Wellcome Trust [reference no. 107768/Z/15/Z] and the UK Foreign, Commonwealth & Development Office, with support from the Developing Excellence in Leadership, Training and Science in Africa (DELTAS Africa) programme. The statements made and views expressed are solely the responsibility of the Fellow.

## Data Availability

data availed as supporting information file

## Notes

### Competing Interest Statement

The authors have declared no competing interest.

### Author Declarations

This study was approved by the Moi Teaching and Referral Hospital / Moi University Institutional Research Ethics Committee (MTRH/MU IREC) (approval number FAN: 0004291) and obtained research permit from the National Commission for Science, Technology and Innovation (NACOSTI) permit Ref number 839919.

